# Maintenance Anesthetics and Postoperative Delirium in Older Adults Undergoing General Anesthesia: A Systematic Review and Network Meta-analysis of Randomized Trials

**DOI:** 10.1101/2025.09.29.25336681

**Authors:** Tomoaki Miyake, Eiki Kanemaru, Kanako Sasaki, Hisashi Noma, Takahiro Mihara

## Abstract

Postoperative delirium (POD) is a frequent and serious complication in older surgical patients. The optimal maintenance anesthetic to prevent POD remains uncertain due to fragmented and inconsistent evidence. This systematic review and network meta-analysis will compare the effectiveness of remimazolam, propofol, sevoflurane, desflurane, isoflurane, xenon, and ciprofol, and establish an evidence-based ranking to guide clinical practice.

**Methods and Analysis:** We will systematically search CENTRAL, Embase, MEDLINE, Web of Science, and major trial registries without language or date restrictions. Two reviewers will independently screen studies, extract data, and assess risk of bias with the ROBUST-RCT tool. Random-effects network meta-analyses will estimate pooled risk ratio (RR) for dichotomous outcomes and mean difference (MD) or standardized mean difference (SMD) for continuous outcomes. Treatment rankings will be summarized using the surface under the cumulative ranking curve (SUCRA) values or P-scores. Certainty of evidence will be appraised with the Confidence in Network Meta-Analysis (CINeMA) framework.

**Ethics and dissemination:** Ethical approval is not required as only published data will be analyzed. Results will be submitted to a peer-reviewed journal. The review was submitted to PROSPERO on 10 August 2025 and registered on 11 August 2025 (CRD420251118351).

## Introduction Rationale

Postoperative delirium (POD) is common in older surgical patients and is associated with prolonged hospitalization, cognitive decline, and increased mortality.^1–6^ With aging populations requiring more surgical interventions, POD prevention has become a perioperative priority.^2,7^ A key mechanism of POD is a neuroinflammatory response to surgical stress, a reaction particularly prominent in the vulnerable aging brain.^8–10^ This neuroinflammation highlights the choice of anesthetic as a key modifiable factor.^11^ Yet existing evidence across anesthetic agents is inconsistent and lacks a clinically oriented, comparative ranking.

The recent introduction of remimazolam has further complicated anesthetic choices.^12–14^ Remimazolam offers favorable pharmacokinetic properties for older adults, including hemodynamic stability and an ultra-short duration of action.^13,15,16^ However, its classification as a benzodiazepine raises significant concern, as this class is well-established to increase POD risk.^17–20^ Critically, no synthesis has positioned remimazolam within the comparative effectiveness landscape of major maintenance anesthetics for POD in older adults.

## Objective

This systematic review and network meta-analysis aimed to compare maintenance anesthetics for their association with POD in older adults and to generate comparative treatment rankings with appropriate assessment of transitivity, consistency, and certainty of evidence.

## Methods Study design

This protocol describes the methods for a systematic review and network meta-analysis of randomized controlled trials. It will be conducted in accordance with the Cochrane Handbook for Systematic Reviews of Interventions (version 6.4) ^21^. The protocol adheres to the Preferred Reporting Items for Systematic Review and Meta-Analysis Protocols (PRISMA-P)^22^ statement, and the final review will follow the PRISMA extension for Network Meta-Analyses (PRISMA-NMA).^23^ This protocol has been registered in the International Prospective Register of Systematic Reviews (PROSPERO; CRD420251118351).

## Eligibility Criteria

Only randomized controlled trials (RCTs) will be eligible for inclusion. Secondary analyses of RCTs will be included if they report relevant outcomes. All other study designs, including quasi-randomized, non-randomized, and observational studies, will be excluded.

## Population

We will include studies enrolling older adults undergoing surgical procedures under general anesthesia. The definition of “older adults” will follow that of each individual study.

## Interventions

Maintenance of general anesthesia with one of the following agents: propofol, remimazolam, ciprofol, sevoflurane, desflurane, isoflurane, or xenon.

## Comparators

Any direct, head-to-head comparison between the eligible maintenance anesthetics.

## Outcomes

### Primary outcome

The primary outcome will be the incidence of postoperative delirium, diagnosed using one of the following validated tools or criteria:

- Confusion Assessment Method (CAM)^24^
- CAM-ICU^25^
- 3-Minute Diagnostic Interview for CAM (3D-CAM)^26^
- Intensive Care Delirium Screening Checklist (ICDSC)^27^
- Nursing Delirium Screening Scale (Nu-DESC)^28^
- Delirium Rating Scale-Revised-98 (DRS-R-98)^29^
- Diagnostic and Statistical Manual of Mental Disorders (DSM) criteria^30^

Assessment windows will be accepted as defined in each trial. When multiple postoperative time points or cumulative incidence estimates are reported within a trial, the earliest eligible postoperative window will be extracted to avoid double-counting. If a single trial reports POD incidence using more than one validated instrument, we will extract the result from the instrument with the highest priority according to the prespecified hierarchy below. The hierarchy is informed by diagnostic-accuracy evidence and international guidance.

Non-ICU settings (wards, PACU once extubated). Highest to lowest priority:

1. DSM
2. CAM or 3D-CAM
3. DRS-R-98
4. Nu-DESC

ICU setting. Highest to lowest priority:

1. DSM
2. CAM-ICU
3. ICDSC
4. DRS-R-98
5. Nu-DESC

Secondary outcomes:

- Intraoperative hypotension, defined as per the included studies and reported as per-patient incidence.
- Emergence agitation, defined as reported by trial authors when assessed during emergence or in the post-anesthesia care unit and reported as per-patient incidence, including scales such as the Richmond Agitation-Sedation Scale (RASS) or the Sedation-Agitation Scale (SAS).
- Delayed neurocognitive recovery within 30 days, assessed using cognitive screening tools such as the Mini-Mental State Examination (MMSE) or the Montreal Cognitive Assessment (MoCA). When multiple postoperative cognitive assessments are available, the earliest eligible assessment on or after postoperative day 3 and within 30 days will be used.
- Thirty-day all-cause mortality.

## Information Sources

We will search MEDLINE (via PubMed), Embase, the Cochrane Central Register of Controlled Trials (CENTRAL), and Web of Science. In addition, we will search trial registries including ClinicalTrials.gov, the WHO International Clinical Trials Registry Platform (ICTRP), the EU Clinical Trials Register (EUCTR), the University Hospital Medical Information Network Clinical Trials Registry (UMIN-CTR), and the Japan Registry of Clinical Trials (jRCT). We will also screen reference lists of included studies and relevant systematic reviews. The initial searches covered records from inception to 13 August 2025. We plan to rerun all four database searches from inception on 13 October 2026 using revised strategies that include isoflurane-specific terms. No language restrictions will be applied.

## Search Strategy

We will develop database-specific search strategies using both controlled vocabulary (e.g. MeSH) and free-text terms, covering concepts for older adults, surgical procedures, and maintenance anesthetics (propofol, remimazolam, ciprofol, sevoflurane, desflurane, isoflurane, xenon). The revised database-specific search strategies are presented in Tables 1a–1d.

**Table 1a.**
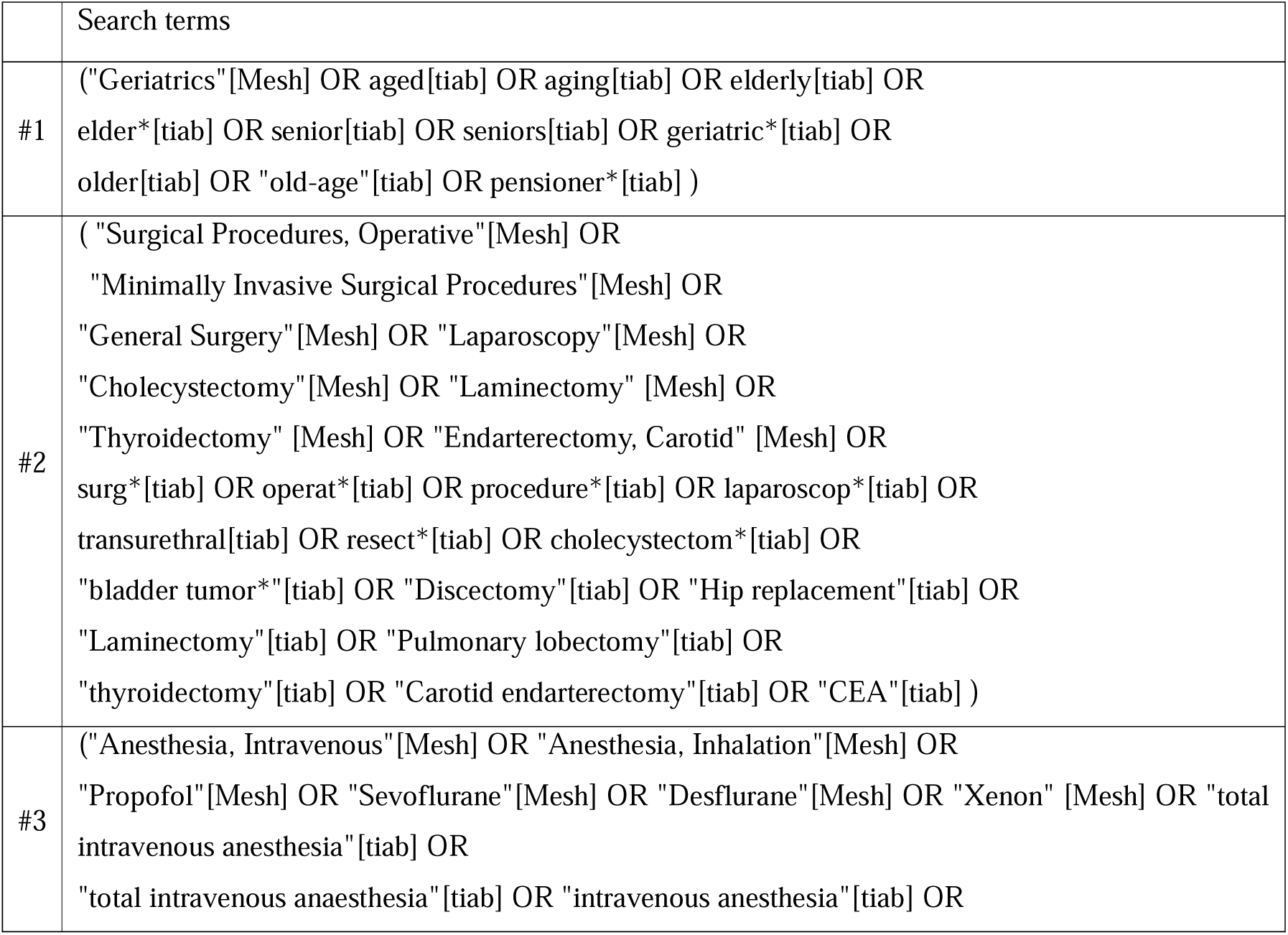
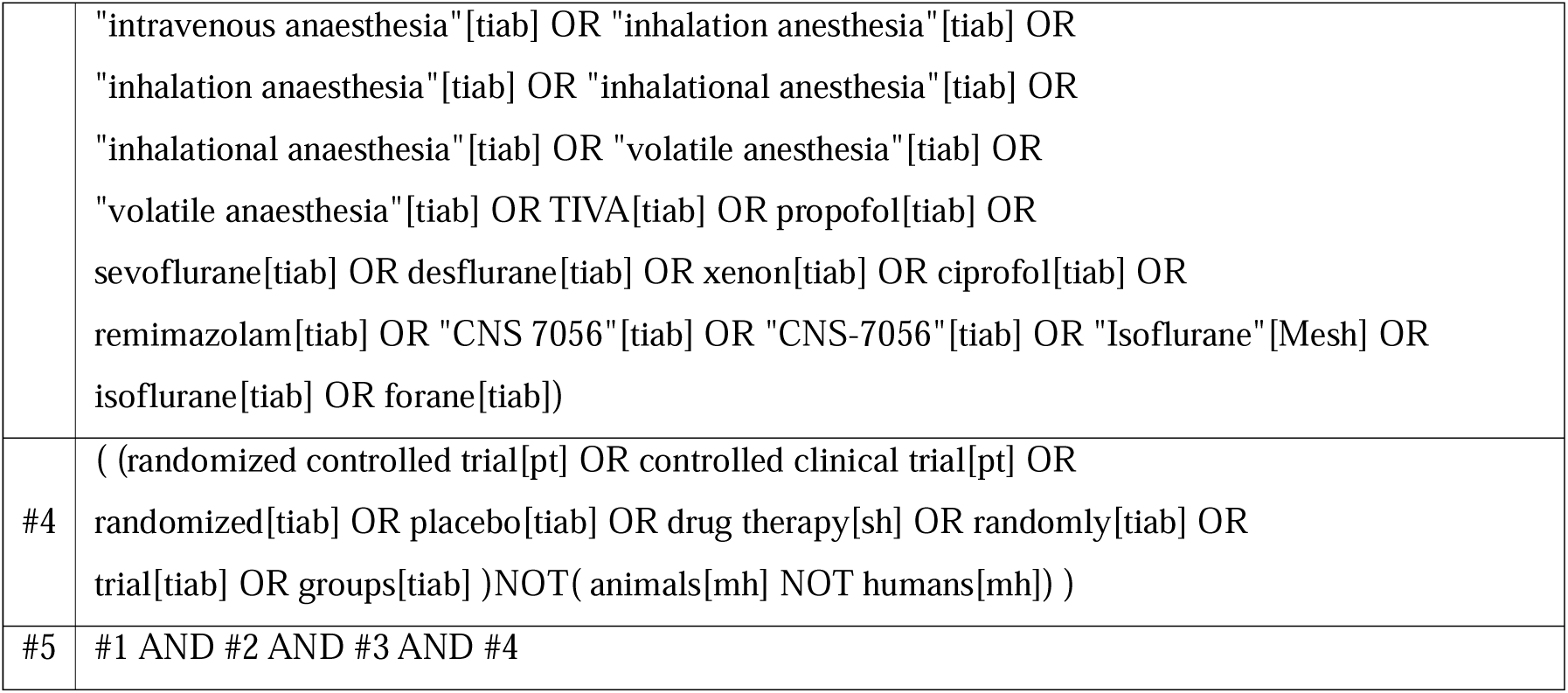
Search strategy for PubMed

**Table 1b.**
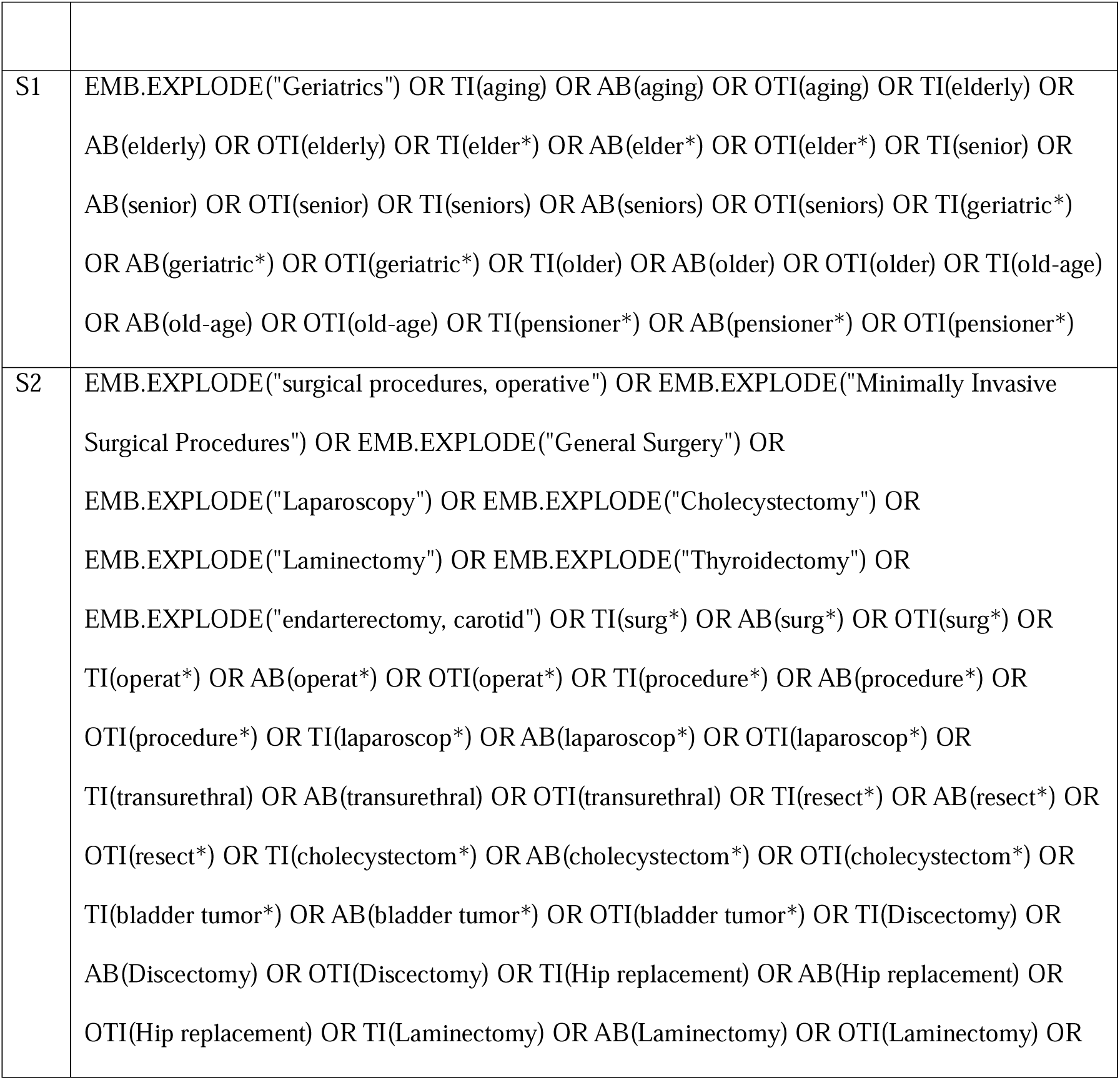
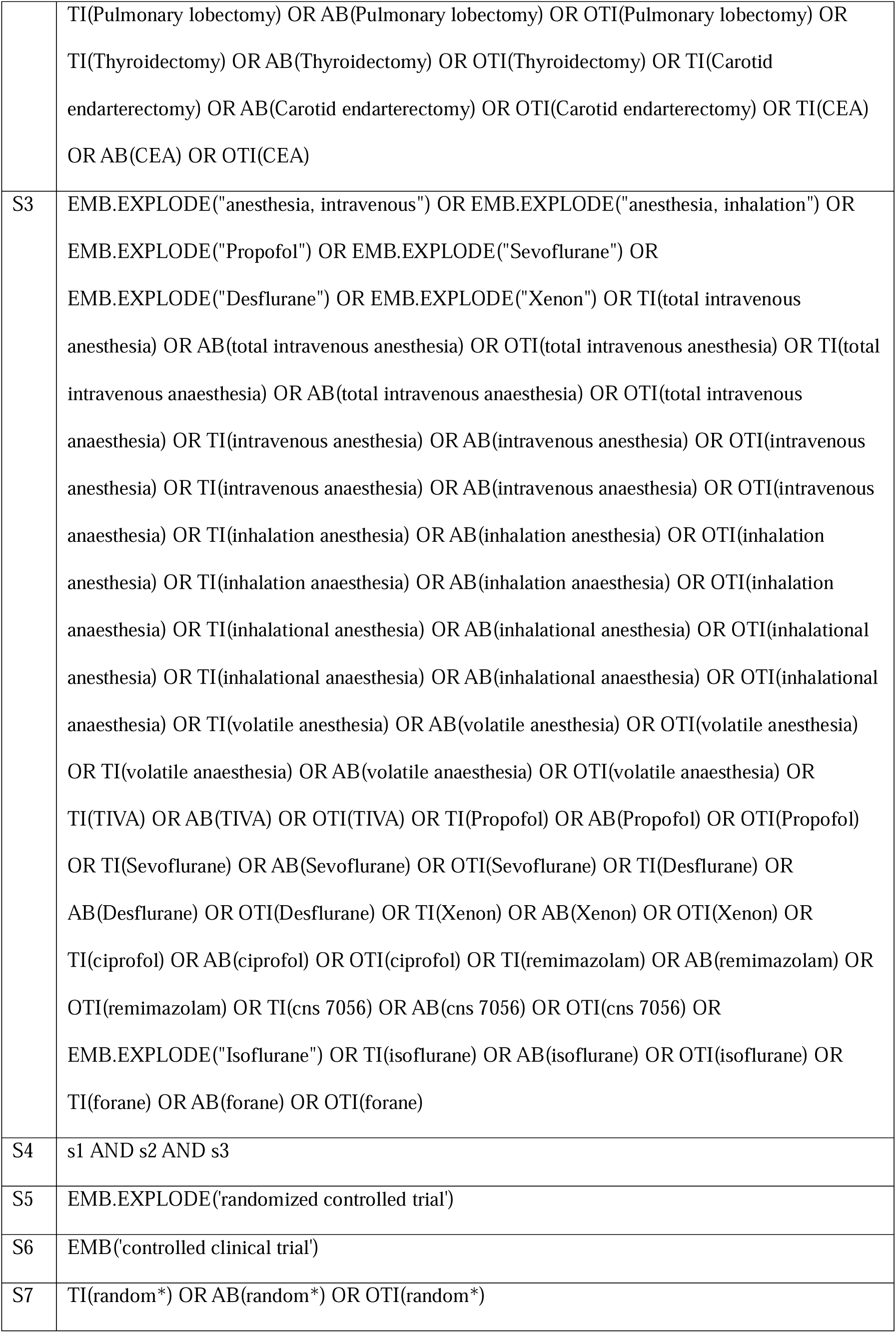
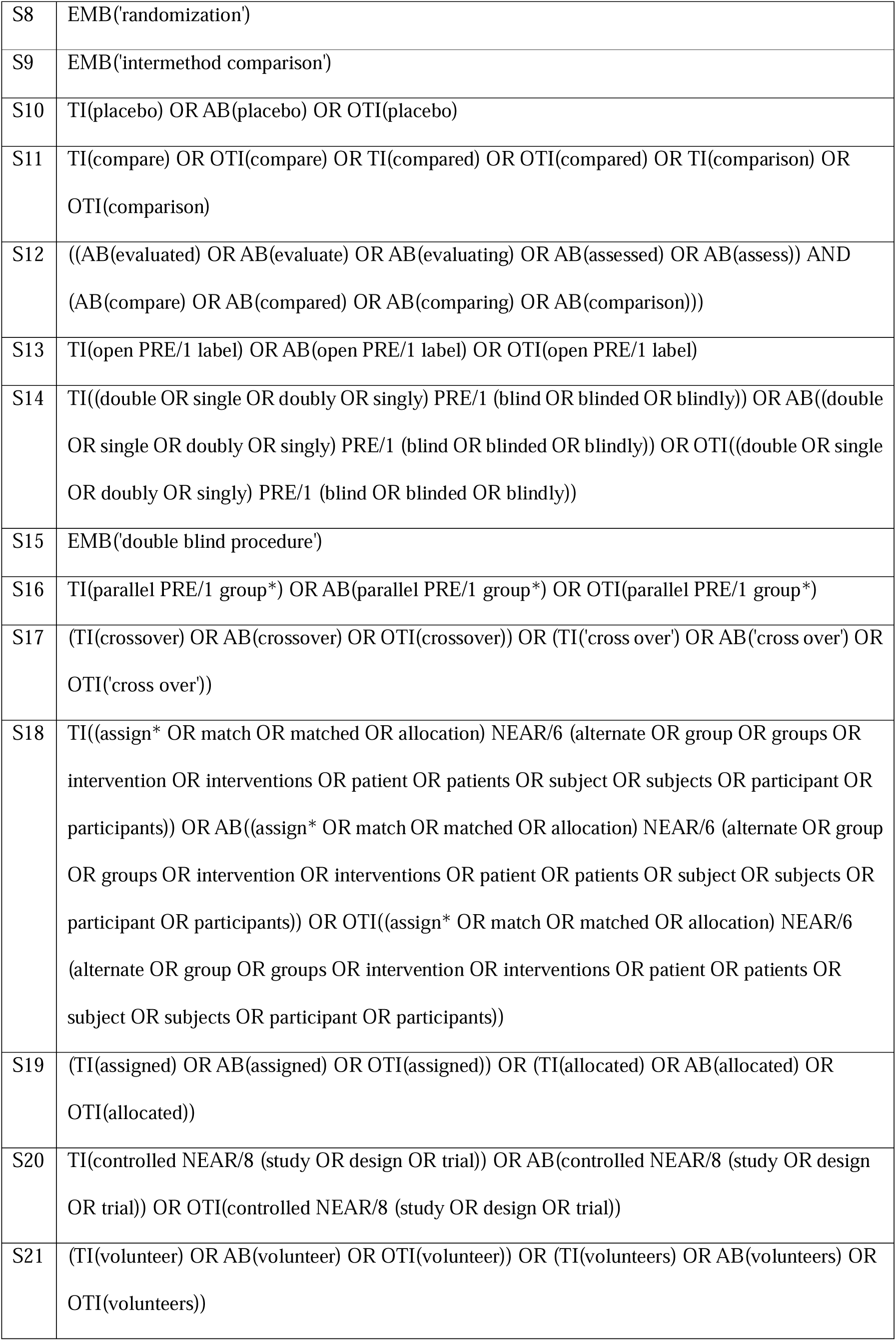
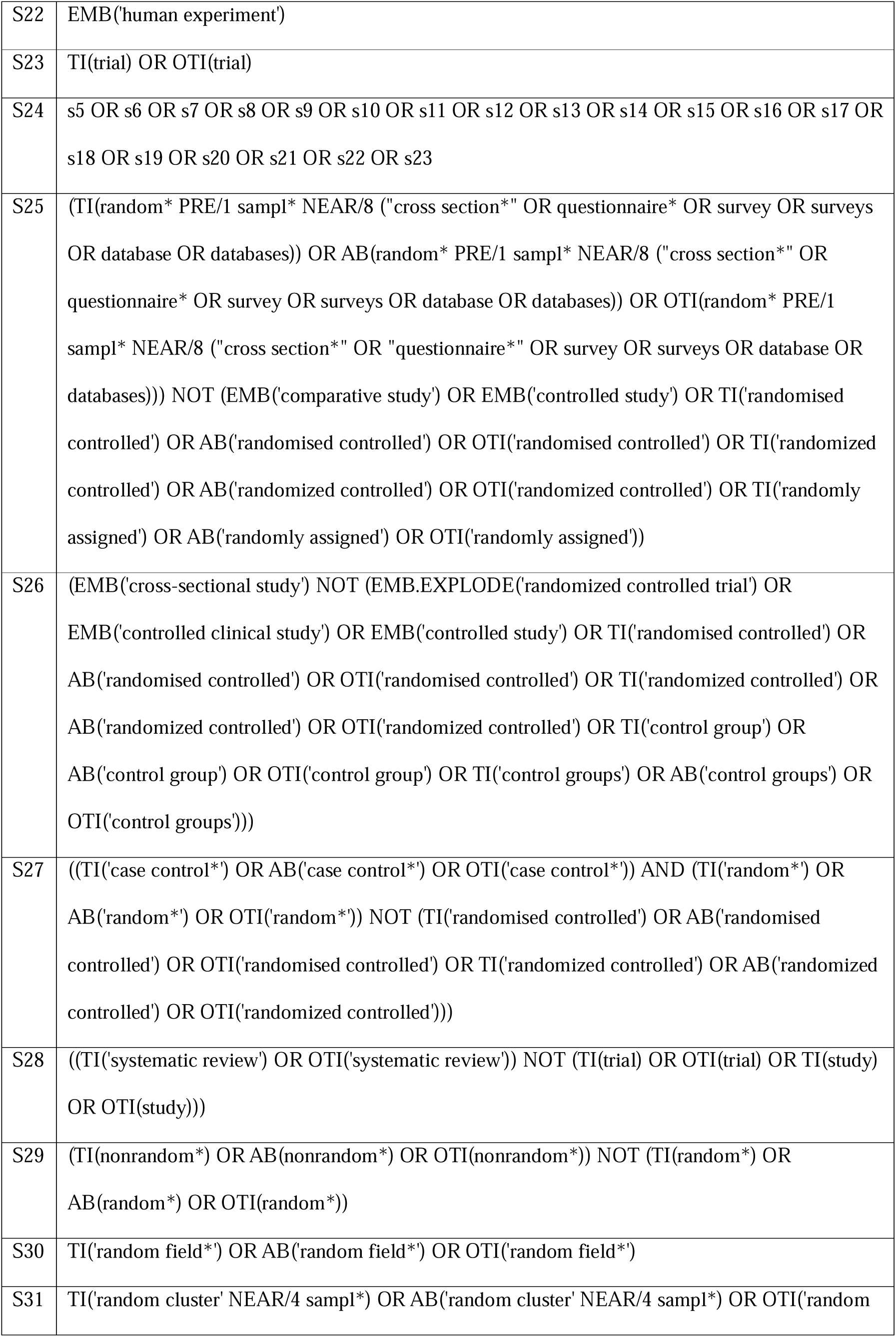
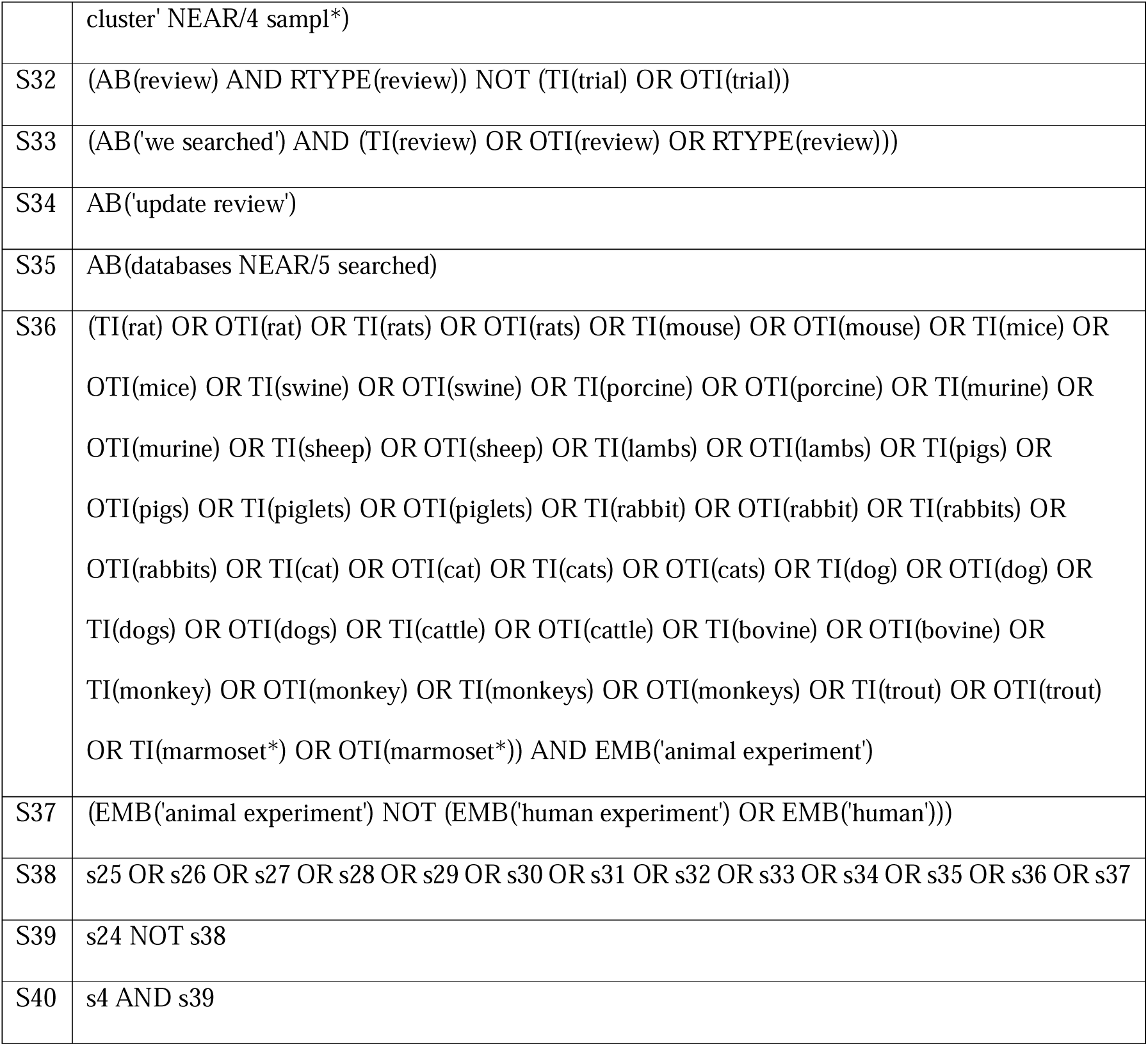
Search strategy for Embase.

**Table 1c.**
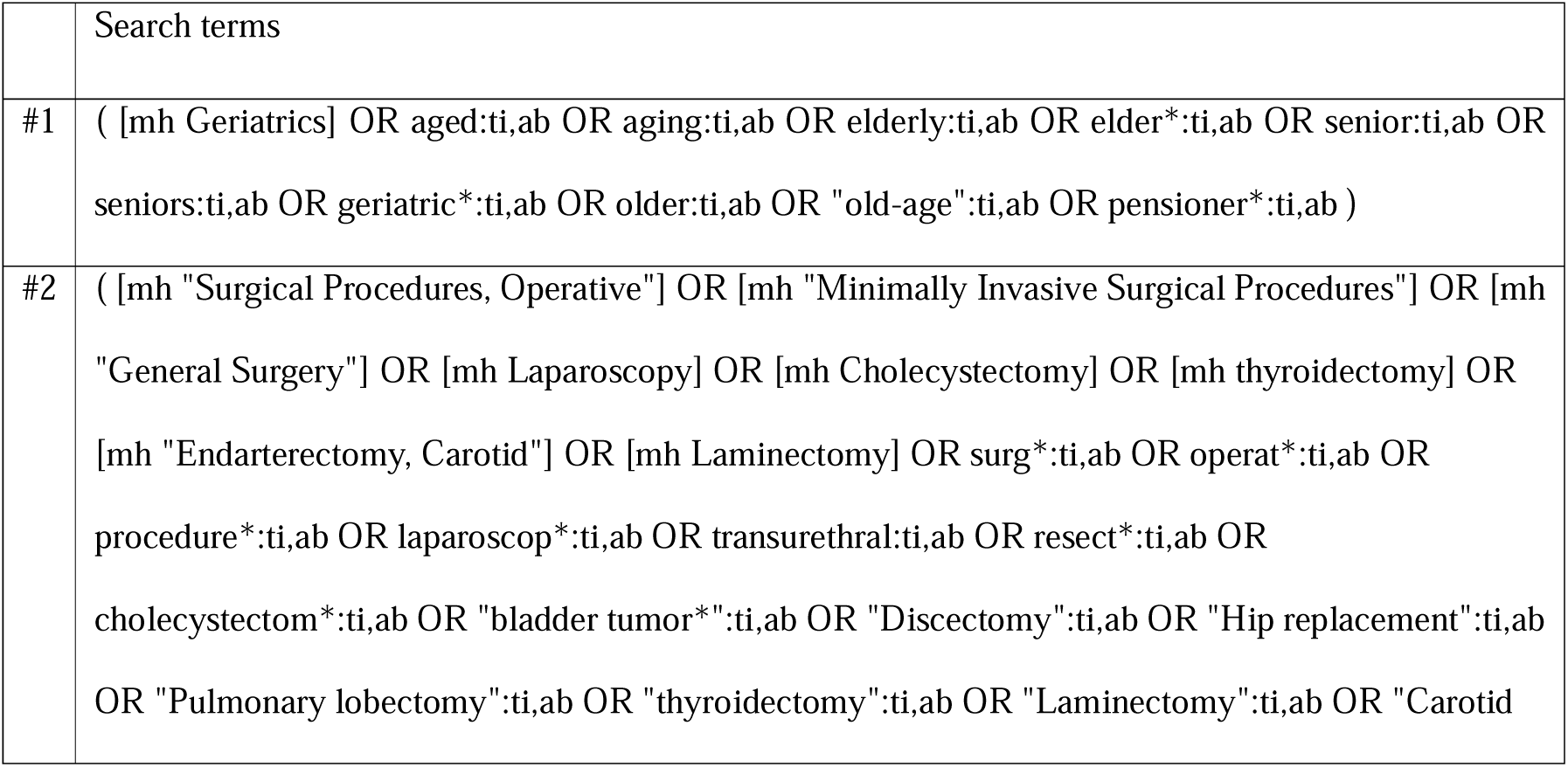
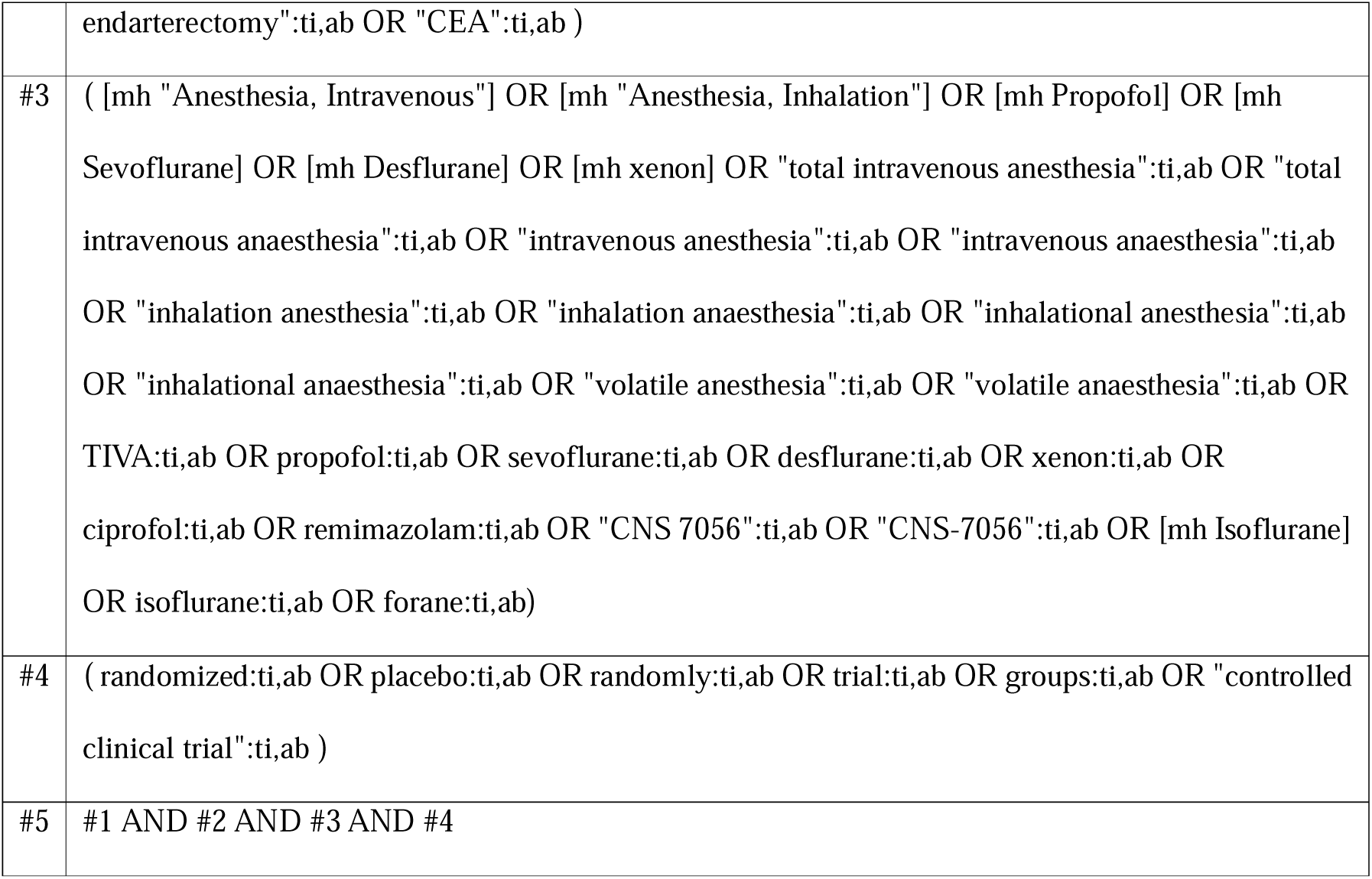
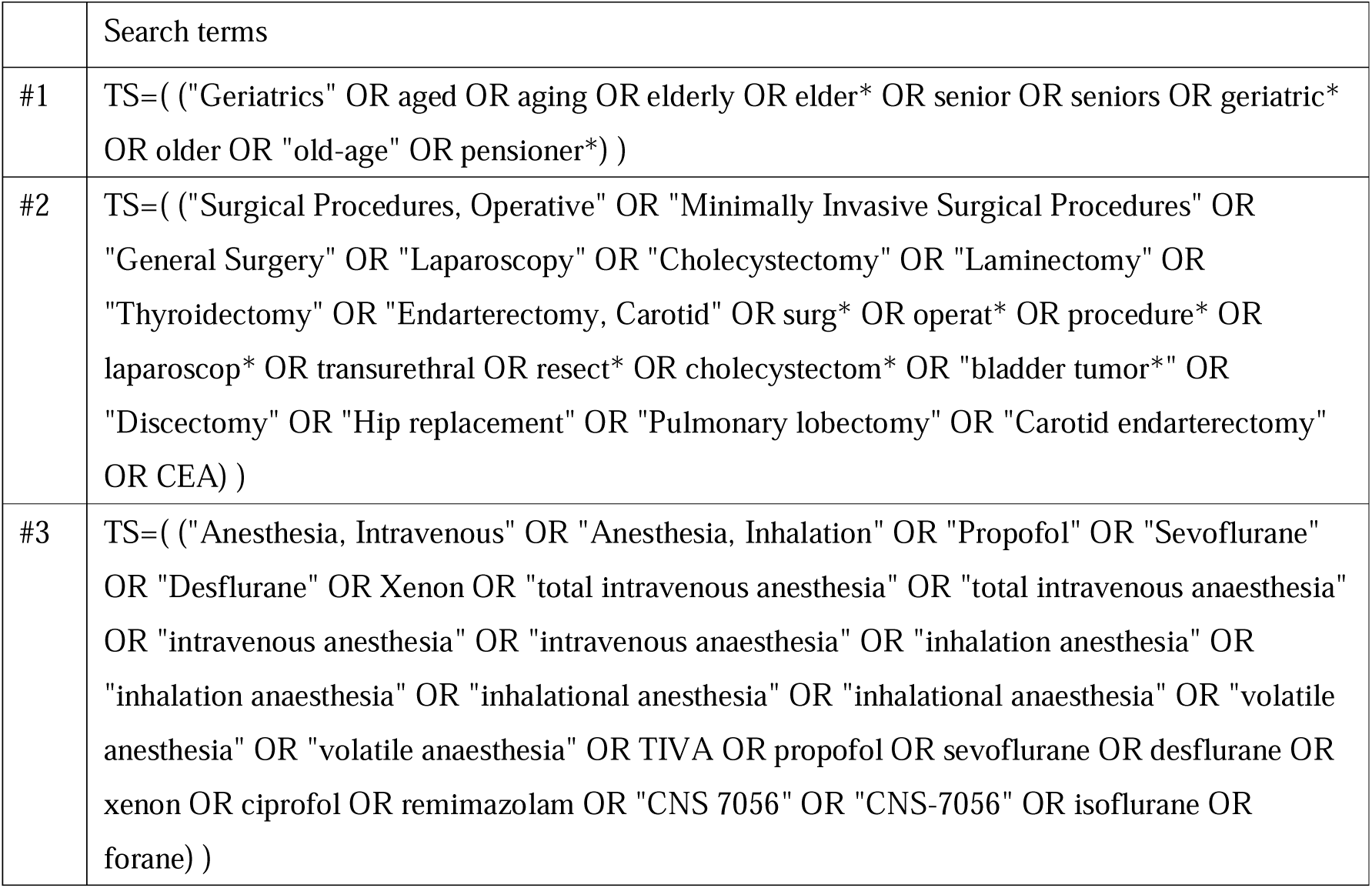
Search strategy for CENTRAL.

**Table 1d.**
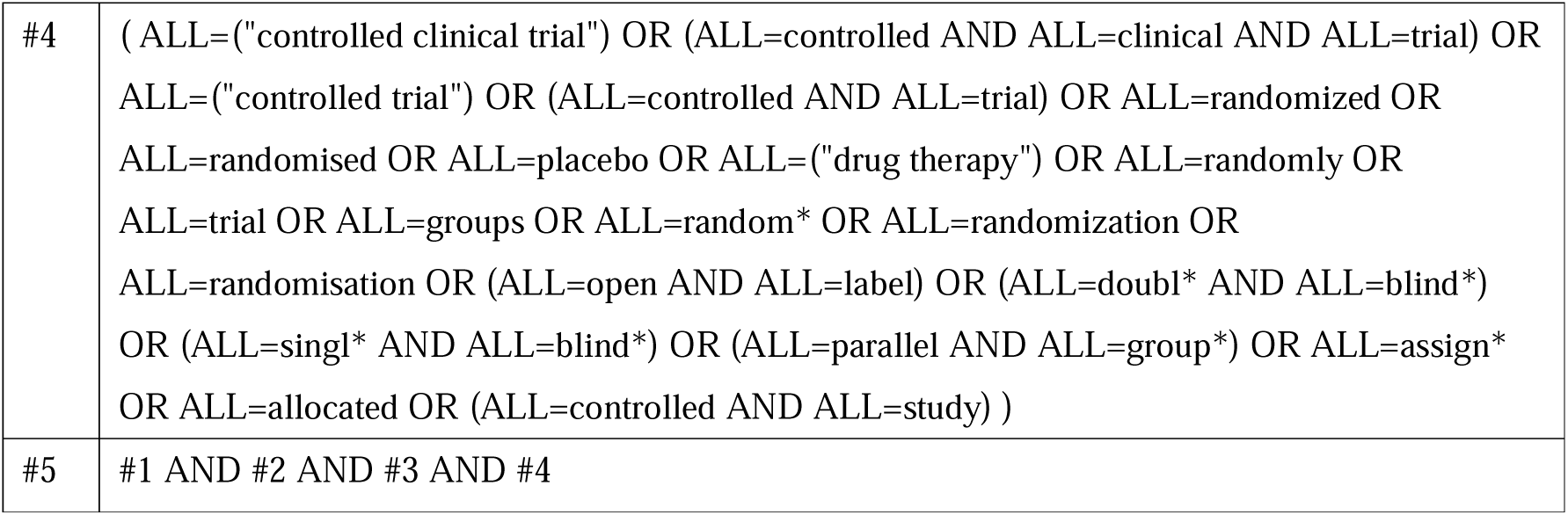
Search strategy for Web of Science

### Data management

Citations retrieved from each database will be exported in RIS format and imported into Covidence (Veritas Health Innovation, Melbourne, Australia), a web-based platform for systematic review management. Covidence will be used to de-duplicate records and to facilitate independent screening of titles and abstracts by multiple reviewers.

## Study Selection

To ensure duplicate independent screening, one reviewer (T. Miyake) will screen all titles and abstracts, while two other reviewers (E.K., K.S.) will each independently screen half of the records. The same process will be applied to full-text review. Discrepancies will be resolved by discussion, with arbitration by a senior author (T. Mihara) if necessary.

## Data Extraction

Two reviewers will independently extract data from all included studies using a standardized extraction form.

Data items will include:

- **Study characteristics:** first author, publication year, country, number of centers, trial design, registry/identifier, sample size, funding source, and conflicts of interest.
- **Participants:** age, sex, type of surgery, inclusion/exclusion criteria, American Society of Anesthesiologists Physical Status classification.
- **Interventions and comparators:** anesthetic agent used for maintenance, dosing regimen or target, induction agent, airway management, and relevant co-interventions (e.g. remifentanil).
- **Outcomes:** for each prespecified outcome (postoperative delirium, intraoperative hypotension, emergence agitation, delayed neurocognitive recovery within 30 days, 30-day all-cause mortality), we will extract the definition used, diagnostic/assessment tools, assessment frequency and time window, numbers randomized and analyzed per arm, and numerical data (event counts and denominators for dichotomous outcomes; means and standard deviations for continuous outcomes).

## Risk of Bias Assessment

We will assess risk of bias using the Risk Of Bias instrument for Use in SysTematic reviews-for Randomised Controlled Trials (ROBUST-RCT).^31^ Two reviewers will evaluate each trial independently, with disagreements resolved by discussion and, if necessary, adjudication by a third reviewer. ROBUST-RCT applies a two-step approach. Step 1 documents what actually occurred in the trial, while Step 2 provides an overall judgment of risk of bias (Definitely/Probably low vs Probably/Definitely high), without mechanically deriving it from Step 1.

We will evaluate the six core domains specified by ROBUST-RCT:

- Item 1 — Random sequence generation
- Item 2 — Allocation concealment
- Item 3 — Blinding of participants
- Item 4 — Blinding of healthcare providers
- Item 5 — Blinding of outcome assessors
- Item 6 — Outcome data not included in analysis

Optional domains will be recorded when relevant (e.g. baseline balance, co-interventions, selective reporting, early stopping).

We will derive an overall study-level rating from the Step 2 judgments as follows:

- Low risk — all six core domains are Definitely low or Probably low.
- High risk — at least one core domain is Definitely high, or two or more are Probably high.
- Some concerns — all other combinations.

## Data Synthesis

### Summary measures (measures of treatment effect)

For dichotomous outcomes, we will use risk ratios (RRs) with 95% confidence intervals. For continuous outcomes, we will use mean differences (MDs) when scales are identical and standardized mean differences (SMDs) when different scales are used, each with 95% confidence intervals (CIs).

## Pairwise meta-analysis

Where sufficient trials allow direct head-to-head comparisons between eligible maintenance anesthetics, we will perform pairwise meta-analyses using a random-effects model. Risk ratios will be used for dichotomous outcomes and mean differences or standardized mean differences for continuous outcomes, each with 95% CIs. Between-study heterogeneity will be quantified with the I² statistic. Multi-arm trials will be analyzed accounting for within-study correlations to avoid double-counting.

## Network meta-analysis

### Review of network geometry

We will construct a network diagram and evaluate the network geometry.^23^ Evaluating the geometry of the network allows for an assessment of the feasibility of NMA, such as determining whether the network of interventions is connected. Additionally, this assessment includes the identification of closed loops of treatments within the network, facilitating the evaluation of inconsistency that is the disagreement between effects estimated from direct and indirect sources.

### Transitivity and inconsistency in NMA

We will statistically evaluate both local and global inconsistency. The local assessment will be performed using the node-splitting method^32^ while the global assessment will be conducted via the design-by-treatment interaction model.^33^ We will perform a random-effects NMA assuming a common between-studies variance across the whole network, with τ² estimated using restricted maximum likelihood (REML). Summary effect measures such as RR will be estimated along with 95% CI. The results of the estimation will be presented using the league table and the Surface Under the Cumulative Ranking curve (SUCRA)^34^, or the P-score, a frequentist version of SUCRA.^35^ We will use the R package “nma” (cran.r-project.org/web/packages/NMA/index.html),^36^ and “netmeta” (cran.r-project.org/web/packages/netmeta/index.html).^37^

## Narrative synthesis

If substantial heterogeneity or an insufficient number of studies prevents quantitative synthesis, we will conduct a systematic narrative synthesis. We will summarize and tabulate the key characteristics and findings of included studies to highlight patterns, similarities, and differences across trials.

## Additional analysis

### Subgroup analysis

Where data permit, we will perform the following a priori subgroup analyses:

- Age: (i) study level meta regression with mean/median age as a covariate, and (ii) subgroup analysis stratified at ≥75 years vs <75 years.
- Type of surgery / surgical complexity: stratification by short duration/minor procedures vs long duration/major procedures, using the definitions or operative times reported within each trial.
- Postoperative care setting: stratification by ICU admission vs general ward return after surgery.

Effect modification will be evaluated by testing subgroup–treatment interaction terms (where supported by the chosen synthesis framework); results will be presented as subgroup specific estimates with interaction P values.

### Sensitivity analysis

To assess robustness, we will conduct:

- Instrument restricted sensitivity (diagnosis of POD): restrict the primary outcome to trials that used CAM family instruments (CAM, CAM ICU, 3D CAM) or the ICDSC, which are widely validated and guideline endorsed for perioperative/ICU populations. This minimizes heterogeneity due to measurement non equivalence across tools.
- Risk of bias restriction: exclude trials judged high risk of bias at the study level under

ROBUST RCT Step 2.

- Older-old population restriction: restrict to trials in which the mean or median age of the analyzed cohort is ≥75 years.

### Small study effects

We will investigate potential small-study effects and publication bias. We will use comparison-adjusted funnel plots and regression-based asymmetry tests to explore small-study effects. In addition, we will assess risk of bias due to missing evidence in the network using the ROB-MEN framework.

## Confidence in Evidence

We will assess the certainty of evidence using the CINeMA (Confidence in Network Meta-Analysis) framework. Two reviewers will perform independent assessments, with disagreements resolved by discussion and, if necessary, arbitration by a third reviewer. CINeMA evaluates six domains: within-study bias, across-studies bias, indirectness, imprecision, heterogeneity, and incoherence. Each comparison will be rated as high, moderate, low, or very low certainty. The judgments will be summarized in summary of findings tables.

## Data Availability

All data produced in the present study are available upon reasonable request to the authors.

## Ethics Approval

Not required. This protocol involves secondary analysis of published or registered data and does not include human participants or identifiable personal data.

## Data and Code Availability

No original data will be generated. Full search strategies and the PRISMA-P checklist will be provided as supplementary materials.

## Version history

- 30 September 2025 (v1): Initial protocol publication.
- 26 July 2026 (v2): Revised outcome definitions and assessment-selection rules after data extraction and before quantitative synthesis. Updated authorship and terminology.
- 13 October 2026 (v3): Corrected the reported PROSPERO submission date and added this version history. Added isoflurane-specific search terms and a plan to rerun all four database searches from inception on 13 October 2026. Updated affiliations.

